# Efficient Chest X-Ray Feature Extraction and Feature Fusion for Pneumonia Detection Using Lightweight Pretrained Deep Learning Models

**DOI:** 10.1101/2025.06.28.25330459

**Authors:** Yashvi Chandola, Vivek Uniyal, Yamini Bachheti

## Abstract

Pneumonia is a respiratory condition characterized by inflammation of the alveolar sacs in the lungs, which disrupts normal oxygen exchange. This disease disproportionately impacts vulnerable populations, including young children (under five years of age) and elderly individuals (over 65 years), primarily due to their compromised immune systems. The mortality rate associated with pneumonia remains alarmingly high, particularly in low-resource settings where healthcare access is limited. Although effective prevention strategies exist, pneumonia continues to claim the lives of approximately one million children each year, earning its reputation as a "silent killer." Globally, an estimated 500 million cases are documented annually, underscoring its widespread public health burden. This study explores the design and evaluation of the CNN-based Computer-Aided Diagnostic (CAD) systems with an aim of carrying out competent as well as resourceful classification and categorization of chest radiographs into binary classes (Normal, Pneumonia). An augmented Kaggle dataset of 18,200 chest radiographs, split between normal and pneumonia cases, was utilized. This study conducts a series of experiments to evaluate lightweight CNN models—ShuffleNet, NASNet-Mobile, and EfficientNet-b0—using transfer learning that achieved accuracy of 90%, 88% and 89%, prompting the task for deep feature extraction from each of the networks and applying feature fusion to further pair it with SVM classifier and XGBoost classifier, achieving an accuracy of 97% and 98% resepectively. The proposed research emphasizes the crucial role of CAD systems in advancing radiological diagnostics, delivering effective solutions to aid radiologists in distinguishing between diagnoses by applying feature fusion, feature selection along with various machine learning algorithms and deep learning architectures.

## 1. Introduction

Pneumonia is an inflammatory respiratory condition triggered by pathogenic microorganisms, including bacterial agents like Streptococcus pneumoniae, viral particles, or, in rare instances, fungal or parasitic organisms. The infection causes fluid accumulation in the alveoli, impairing gas exchange and leading to respiratory distress. While it affects all age groups, immunocompromised populations— notably children under five years old and elderly adults over 65—face the highest mortality risk. Global health data reveals alarming statistics, with approximately 15 fatalities per 100,000 individuals annually, predominantly in low-resource settings where pediatric cases account for nearly one million preventable deaths each year. With over half a billion reported cases worldwide, pneumonia has earned the grim moniker "the silent killer" due to its insidious progression and diagnostic challenges. Although no universal cure exists, preventive measures like pneumococcal conjugate vaccines (PCV) and Haemophilus influenzae type b (Hib) immunizations have significantly reduced incidence rates. Despite these advances, lower respiratory infections rank as the third-leading cause of death globally, with pneumonia representing the most severe subset. Accurate diagnosis traditionally relies on chest X-ray analysis, yet even experienced radiologists may struggle to differentiate bacterial from viral etiologies, potentially delaying appropriate treatment. In resource-constrained regions, the scarcity of specialized healthcare professionals underscores the critical need for computer-aided detection (CAD) systems. Such technologies can augment diagnostic precision, facilitate early intervention, and ultimately reduce mortality rates by supporting clinical decision-making. [1–3].

Deep learning is an advanced subset of artificial intelligence and machine learning that utilizes complex neural networks with multiple processing layers. These architectures are designed to replicate human cognitive functions by creating hierarchical data representations. By progressively analyzing raw input data through successive layers, deep learning models can identify and refine intricate patterns, closely mimicking human perceptual and analytical processes. Among these architectures, Convolutional Neural Networks (CNNs) are widely used for tasks like speech recognition and object detection. Typically, deep learning models are structured into input, hidden, and output layers, with each layer responsible for feature extraction and data transformation to support subsequent learning stages.

In healthcare, deep learning plays a pivotal role in medical imaging by automating disease diagnosis through internal structure visualization. Imaging modalities such as X-rays, CT scans, and ultrasound generate vast datasets that can challenge manual interpretation by radiologists. Deep learning streamlines this process, enabling rapid, precise, and scalable image analysis—key advantages for timely diagnosis and improved patient management.

Chest X-rays serve as a critical, non-invasive diagnostic tool for pulmonary and cardiac conditions, including pneumonia. While they involve lower radiation exposure than CT scans (which provide 3D imaging but require higher radiation doses and contrast agents), X-rays remain indispensable for detecting pneumonia-specific features like pulmonary opacities. Deep learning enhances X-ray diagnostics by efficiently processing large imaging datasets, alleviating radiologists’ workloads, and boosting diagnostic precision. This integration of AI and healthcare facilitates earlier disease detection and optimizes clinical outcomes. Pneumonia, in particular, manifests radiologically as localized or lobar pulmonary opacities, which deep learning models can reliably identify.

Figure 1 (a) displays chest radiographs with a clean and transparent lung portion hence representig normal chest, whereas Figure 1.1 (b) shows chest radiographs with clouded airspace or a radio opaque lung section, clearly representing lungs infected with pneuonia.

**Fig. 1.1.**
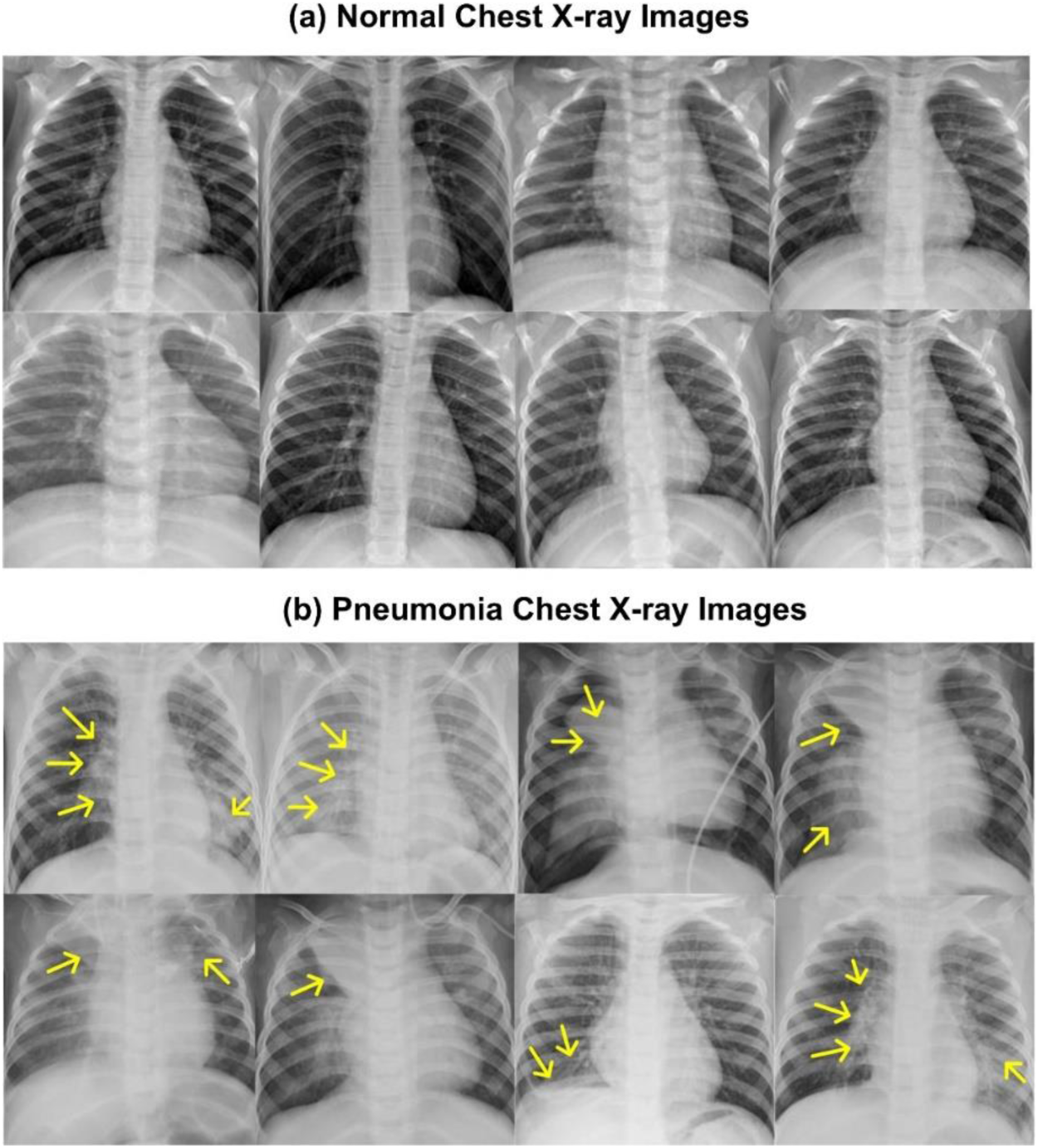
Illustrative represenatation of Chest radiographs (a) A Radiograph showing Normal chest, (b) A Radiograph showing Pneumonia Infected chest.

## 2. Literature Review

The review of many Computer-Aided Diagnostic (CAD) systems established throughout the eons by researchers for 2-class categorization as well as multiple class classification of chest radiographs has been expanded in the following sections.

### 2.1 Computer-Aided Diagnostic (CAD) system designs for Chest Radiographs with focus on Machine Learning based algorithms

Recent advancements in medical image analysis have demonstrated the effectiveness of machine learning techniques for pneumonia detection in chest X-rays. Chandra and Verma (2020) [26] proposed an innovative approach leveraging lung region-of-interest (ROI) segmentation to improve classification accuracy between normal and pneumonia cases. Their comparative analysis revealed that segmentation significantly enhanced performance across classifiers, with logistic regression achieving the highest accuracy (95.63%) when using ROI-based processing, compared to 91.50% without segmentation. Other classifiers like multi-layer perceptron (95.38%) and random forest (94.41%) also showed notable improvements with segmentation.

Further supporting these findings, Al Mamlook et al. (2020) [27] reported even higher accuracy rates, with random forest reaching 97.61% and CNN achieving 98.46%, underscoring the potential of deep learning in this domain. Earlier studies by Oliveira et al. (2008) [28] and Sousa et al. (2013) [29] explored traditional machine learning methods, with Oliveira employing Haar wavelet transforms for feature extraction in a k-NN classifier, while Sousa compared SVM (77%), k-NN (70%), and Naïve Bayes (68%), later improving results using dimensionality reduction techniques like kernel-PCA. Depeursinge et al. (2010) [30] and Yao et al. (2011) [31] further validated SVM’s utility, reporting accuracies of 88.30% and 80.00%, respectively. Naydenova et al. (2015) [32] advanced this work by integrating feature selection with ensemble methods, achieving 97.80% accuracy using a hybrid random forest-SVM approach.

Recent studies have introduced more sophisticated techniques. For instance, Ahmed et al. (2023) [121] combined EfficientNetV2 with XGBoost, achieving 95.2% accuracy while emphasizing model interpretability through SHAP values. Similarly, Gupta et al. (2024) [124] proposed a lightweight MobileNetV3-XGBoost pipeline for tuberculosis detection, attaining 92.8% accuracy with reduced computational costs—a critical consideration for clinical deployment. Notably, Tseng and Tang (2023) [123] optimized XGBoost with feature selection for brain tumor detection, demonstrating its versatility beyond pulmonary diseases.

While prior research predominantly relied on SVM and basic classifiers. The current study addresses this gap by integrating feature fusion with SVM and XGBoost, combining the strengths of traditional and ensemble methods. This approach not only builds on existing segmentation and feature extraction strategies but also leverages XGBoost’s superior handling of imbalanced medical data, as evidenced by recent hybrid models [121–124].

### 2.2 Computer-Aided Diagnostic (CAD) system designs for Chest Radiographs with focus on Deep Learning based CNN networks

Recent advances in deep learning have revolutionized computer-aided diagnosis (CAD) systems for pulmonary conditions. Zech et al. (2018) [2] pioneered a robust classification framework using DenseNet121 with softmax classifier, validating their approach on a comprehensive multi-institutional dataset from NIH, Mount Sinai, and Indiana University networks. This multicenter study demonstrated exceptional generalization capabilities, setting a benchmark for subsequent research. Parallel work by Mubarok et al. (2019) [7] developed an assistive diagnostic system achieving 85.60% accuracy with ResNet50, though Mask R-CNN showed slightly lower performance (78.06%).

Contemporary studies have pushed accuracy boundaries further. Rahman et al. (2020) [8] conducted extensive evaluations across four architectures, with DenseNet201 (98.00%) outperforming ResNet18 (96.40%) and SqueezeNet (96.10%). These findings were corroborated by Elasnaoui and Chawki (2020a) [15], whose comparative analysis of nine architectures revealed MobileNetV2 (96.27%) and ResNet50 (96.61%) as top performers. The field has particularly benefited from benchmark datasets like Kermany et al. (2018) [11]’s Kaggle collection (5,856 images), which enabled their InceptionV3 model to achieve 92.08% accuracy.

Recent innovations have introduced novel architectural modifications. Liang and Zeng (2020) [20] addressed critical overfitting challenges through residual structures and dilated convolutions, while Togacar et al. (2019) [4] demonstrated exceptional performance (99.41% accuracy) combining mRMR feature selection with LDA classification. The trend toward lightweight models is evident in Chandola et al. (2021) [103]’s work, where MobileNetV2 achieved 94% accuracy, improving to 95% with decision fusion.

While existing studies show promise, three key limitations emerge, most systems focus on binary classification that have limited exploration of feature fusion techniques. Khan et al. (2022) [126] developed a triple-attention CNN with 98.6% accuracy for pediatric pneumonia. Our current work introduces novel feature fusion mechanisms combining radiomic features with deep learning embeddings. The evolution from basic CNNs to sophisticated hybrid systems demonstrates remarkable progress, yet opportunities remain for improving generalizability across diverse populations and clinical settings

## 3. Proposed CAD System Design for Chest Radiographs

The proposed Computer-Aided Diagnostic (CAD) system for pneumonia detection in chest X-rays employs a multi-stage pipeline beginning with dataset acquisition, resizing, and augmentation to ensure robust model training. It explores two complementary approaches:

- Lightweight end-to-end CNNs (ShuffleNet, NASNet-Mobile, EfficientNet-b0) CAD system design
- Hybrid CNN-ML CAD system designs where pre-trained networks (ShuffleNet, NASNet-Mobile, EfficientNet-b0) extract features for classifiers like SVM and XGBoost

The proposed work involves a sequence of experiments that also involve uniquely integrating feature across multiple models (ShuffleNet, NASNet-Mobile, EfficientNet-b0) to enhance reliability, while SVM and XGBoost visualizations provide interpretability for clinical trust. Designed for both high accuracy and scalability, it addresses key gaps in existing systems—balancing computational efficiency (critical for low-resource settings) with diagnostic precision, ultimately aiming to reduce radiologist workload and improve early pneumonia detection through automated, explainable AI. The Fig. 3.1 shows the Illustrative representation of the flow of the proposed work.

**Fig. 3.1.**
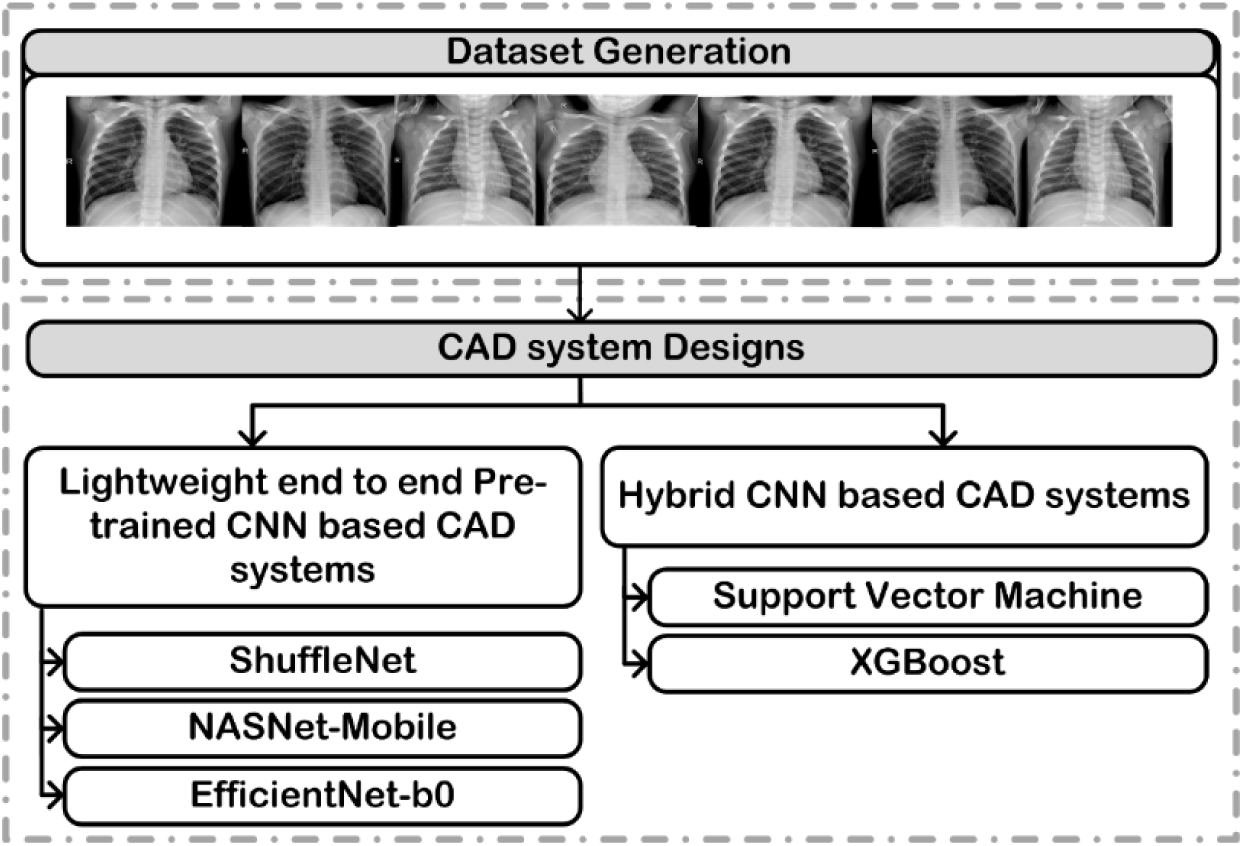
Illustrative Representation of the flow of Proposed work.

### 3.1 Dataset Description

The experimental dataset for this study was obtained from the publicly available Kaggle repository originally curated by Kermany et al. (2018) [38,42]. For balanced experimental evaluation, we utilized a subset of 200 annotated chest X-ray images, with precisely 100 samples representing normal pulmonary cases and 100 demonstrating radiographic evidence of pneumonia. This carefully selected 1:1 class ratio ensures equitable representation of both diagnostic categories in our analytical framework. The dataset’s public accessibility facilitates reproducibility while its clinically validated annotations provide reliable ground truth for model training and validation. For the proposed work the images are downsized while maintaining their aspect ratio in order to maintain the lung form. Both normal and pneumonia chest radiographs are subjected to the identical enhancement strategies in this investigation. The training dataset is expanded to 18,100 radiographs from its original 100 images. First, each image undergoes translation and rotation following a horizontal and vertical flip of these translated and rotated radiographs. A total of 181 images are created by augmenting each image. The research employs a systematic data partitioning strategy, segmenting the complete dataset into three mutually exclusive groups: training, validation, and test sets. The division process begins with random allocation of 50% of the original images to the test set. The remaining half is then subjected to augmentation procedures designed to maintain balanced representation between normal and pathological cases. From this augmented pool, a stratified 10% sample is extracted to serve as the validation set. Notably, both original images and their augmented counterparts are consistently assigned to identical subsets throughout this partitioning process, preserving data integrity and preventing information leakage between sets.

### 3.2 Lightweight Deep Learning based CNN Networks

In modern medicine, Computer-Aided Diagnostic (CAD) systems utilizing Convolutional Neural Networks (CNNs) have become essential, particularly in medical imaging analysis [28–40]. CNNs employ a structured, multi-layered design to identify complex patterns within visual data. These layers—comprising Convolution, Activation, Pooling, Fully Connected, and Softmax—use specialized filters to detect spatial or temporal relationships in images [52–54]. CNNs acquire the ability to recognize local or global features that aid in picture classification during training. To achieve optimal performance in specific tasks, CNNs can be trained effectively using various approaches. One such method is transfer learning, where pre-trained weights from a base network (initially trained on a source dataset) are applied to a target network, enabling quicker adaptation to a new dataset. Another technique is fine-tuning, which involves modifying the parameters of a pre-trained model to suit a different dataset. However, this approach is less frequently utilized in medical image analysis because of the limited availability of extensive datasets. Additionally, training the CNN from scratch is another viable option for obtaining tailored results. It can take days or weeks to train a CNN from scratch, especially when dealing with huge datasets. This can be somewhat alleviated using pre-trained networks, trained on huge benchmark datasets such as ImageNet [25–27]. In proposed work, transfer learning is applied, utilizing a pre-existing lightweight CNN model for the 2-class classification of chest radiographs.

#### 3.2.1 ShuffleNet

ShuffleNet, introduced by Xiangyu Zhang et al., [116] in the paper titled "ShuffleNet: An Extremely Efficient Convolutional Neural Network for Mobile Devices" is precisely understood as a lightweight CNN model specifically designed for mobile devices that lack or have limited resources primarily for computation purposes. The chief goal of ShuffleNet is to achieve high efficiency in regards of the two critical aspects which are namely computational cost (FLOPs) and memory usage, which are essential for mobile platforms. To accomplish this, ShuffleNet uses pointwise group convolution, which progresses by splitting the standard convolution into two convolution operations: depthwise convolution, where each channel that deals with the input is convolved separately, and pointwise convolution, which employs the 1x1 filter that combines the results. The network is designed to balance performance with efficiency, making it apt for real-time applications especially on mobile devices. By improving group convolutions and channel shuffling, ShuffleNet significantly reduces both memory usage and computational requirements, making it a lightweight and fast framework.

#### 3.2.2 NASNet-Mobile

NASNet-Mobile, as given by the researchers Barret Zoph et al. [117], in the paper titled "Learning Transferable Architectures for Scalable Image Recognition" is a CNN architecture designed through NAS with a focus on scalability, efficiency, and transferability across various tasks. NAS is a search process carried out automatically over a large search space to find the most effective architecture by using techniques of reinforcement learning to evolve the neural networks toward performing well for specific tasks. Thus, NASNet-Mobile was designed as a highly efficient architecture for constrained computational resources and memory on a mobile device.

The core notion behind NASNet-Mobile is the use of convolutional layers along with operations that result in maximization of performance while bringing down the computation cost. Such architecture would include a stack of cells since cells are defined as the primary building blocks to be optimized for the best compromise between accuracy and efficiency for a task like image classification. NASNet-Mobile is a condensed version of NASNet architecture developed specifically for a mobile platform. NASNet-Mobile employs reduced kernel sizes with refined operational methods to achieve quite impressive performance for mobile devices endowed with limited computing power.

#### 3.2.3 EfficientNet-b0

EfficientNet-B0, as it is described by Mingxing Tan and Quoc V. Le within the paper titled "EfficientNet: Rethinking Model Scaling for Convolutional Neural Networks" [118], has been developed among the EfficientNet family of models to balance good performance with computing efficiency. EfficiencyNet-B0 is the smallest architecture that provides a baseline scale for other developed EfficientNet variants. The key idea of EfficientNet is the process that scales the dimensions of the network in terms of depth of the network, width of the network, and resolution of a network all at an even rate with one another, thereby improving both at the same time.

Instead of just simply scaling the complexity of the network in terms of enhancing the depth of the network (by adding more layers) or increasing the width (by adding more neurons per layer), as usual in traditional scaling methods, EfficientNet practices the application of a compound scaling method where all three dimensions, namely, depth of the network, width of the network, and resolution of the network, are scaled in such a manner that the network grows in a harmonious manner with the prime aim to maintain optimal balance between the three essential dimensions rather than scaling one in a way to degrade the rest.

### 3.3 Machine Learning Algorithm

#### 3.3.1 Support Vector Machine (SVM)

Support Vector Machines (SVM) represent a powerful supervised learning methodology widely applied to both classification and regression tasks in machine learning. The algorithm’s fundamental principle revolves around constructing an optimal decision boundary that maximizes the separation margin between distinct classes, thereby enhancing generalization performance on unseen data. This maximum-margin classifier operates by identifying support vectors - the most critical data points that define the boundary between classes. For complex, non-linear datasets where linear separation is impossible in the original feature space, SVM employs sophisticated kernel functions to implicitly map the input data into higher-dimensional spaces where linear separation becomes achievable. Commonly used kernel functions include the Radial Basis Function (RBF), polynomial, and sigmoid kernels, each offering unique advantages for different data characteristics [84,103,109]. SVMs demonstrate particular strength in high-dimensional spaces and cases where the number of dimensions exceeds the number of samples, making them well-suited for medical imaging applications. Additionally, their inherent resistance to overfitting, especially when combined with appropriate regularization, contributes to reliable performance across diverse datasets. The algorithm’s mathematical foundation in statistical learning theory provides strong theoretical guarantees about its generalization capabilities, further reinforcing its popularity in both academic research and practical implementations.

#### 3.3.2 XGBoost

XGBoost (eXtreme Gradient Boosting) represents a powerful advancement in machine learning, employing an ensemble approach that builds upon gradient-boosted decision trees [119–121]. This algorithm operates through an iterative process where successive decision trees are constructed to rectify the residual errors of preceding models, progressively refining predictions through gradient descent optimization. A distinctive characteristic of XGBoost lies in its sophisticated regularization framework, incorporating both L1 (Lasso) and L2 (Ridge) penalties to control model complexity and prevent overfitting - a critical feature when working with medical imaging data where generalization is paramount. The implementation efficiently leverages parallel computing capabilities to accelerate model training while maintaining computational resource efficiency. Notably, XGBoost incorporates native functionality to handle missing values intelligently, automatically learning appropriate imputation strategies during the training process. These combined attributes - regularization, parallelization, and robust data handling - render XGBoost particularly effective for processing large-scale datasets while maintaining model interpretability. The algorithm’s inherent scalability allows it to accommodate diverse data types and sizes without compromising performance, making it adaptable to various problem domains. Furthermore, XGBoost includes built-in cross-validation support and early stopping mechanisms, enabling automated optimization of training iterations to maximize predictive accuracy while preventing unnecessary computation. This combination of technical features has established XGBoost as a preferred choice for competitive machine learning applications and real-world implementations where both accuracy and efficiency are prioritized. The algorithm’s flexibility extends to supporting various objective functions and evaluation metrics, allowing customization for specific use cases such as medical diagnosis tasks [120–126].

### 3.4 Feature Extraction

Feature extraction essentially deals with the vital task of extracting and mining substantially noteworthy information from the input image dataset. One major issue is that many variables are needed to effectively describe extracted features in huge datasets, this need of large amount of variable tends to be expensive in terms of both cost and computation time. Learning-based techniques or manual methods can be used for feature extraction. The proposed work focuses on extraction of deep features from the average pool layer of the three (ShuffleNet, NASNet-Mobile, EfficientNet-b0) pre-trained lightweight networks. A total of 544 features from ShuffleNet, 1056 features from NASNet-Mobile and 1280 features from EfficientNet-b0 are extracted from the average pooling layer of each lighweight CNN model.

### 3.5 Feature Fusion

Feature fusion is the process of combining discriminative features extracted from multiple sources such as different neural networks, layers, or modalities to create a more robust, generalized, and informative representation for machine learning tasks such as classification, detection, or segmentation. The common techniques of feature fusion include:

- **Concatenation**: Stacking features end-to-end to preserve all original information.
- **Element-wise Sum/Average**: Merging features by adding or averaging values at each dimension.
- **Attention-based Fusion**: Weighting features dynamically based on their importance for adaptive integration.

The choice of fusion method depends on the task—concatenation for complementary features, sum/average for structurally similar features, and attention mechanisms for prioritizing critical features—with each method offering unique advantages in balancing information retention and model complexity. In proposed work this technique leverages the strengths of different feature extractors, such as the Global Average Pooling (GAP) layers from multiple CNNs like ShuffleNet, NASNet-Mobile, and EfficientNet-b0, to enhance model performance by integrating complementary information. The proposed work deals with deep feature extraction from each model followed by the concatenation based feature fusion to form a ultimate deep feature set of 2880 features.

### 3.6 Feature Selection

The act of distilling an initial big quantity of unprocessed and unanalysed data into a highy controlled, practicable, useful, and smaller subset of features is poularly called as the task of feature selection. These chosen features are frequently many, and managing sizable feature sets necessitates a variety of factors. This is addressed by feature selection, which lowers the number of variables required by limiting the feature set. The keywords Feature selection and feature dimensionality reduction are the two popularly used techiques to describe the these essential tasks of selecting a useful subsection of features from the original set of features or combining features to create new ones that are more useful anf filled with information respectively. Here, the only goal is to create a renewed feature set that accurately reflect the original dataset or is close to the original set as much as possible. Feature selection approaches include two categories of strategies namely, filter-based and wrapper-based strategies. Filter-based techniques are more commonly utilized, including chi-squared tests, box plots, and correlation-based feature selection. Although wrapper-based techniques need a significant amount of computation, model-based genetic algorithms, such as GA-SVM or GA-kNN, are utilized. Among the various feature selection techniques the proposed work focuses on correlation-based feature selection. The deep feature set of a total of 2880 features formed after fetaure fusion further undergoes corelation based feature selection resulting in a reduced feature set of 266 features. The popularly used feature selection techniques are shown in Fig. 3.2.

**Fig. 3.2.**
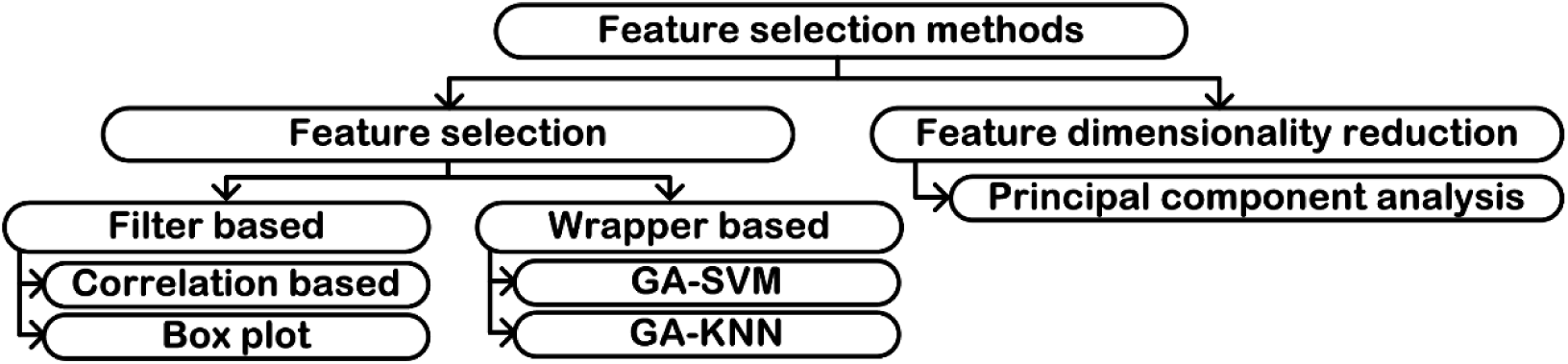
Feature selection methods.

## 4. Experiments

Figure 4.1 Illustrates the experimental procedure pf the proposed work for classifying chest radiographs into normal and pneumonia classes.

**Fig. 4.1.**
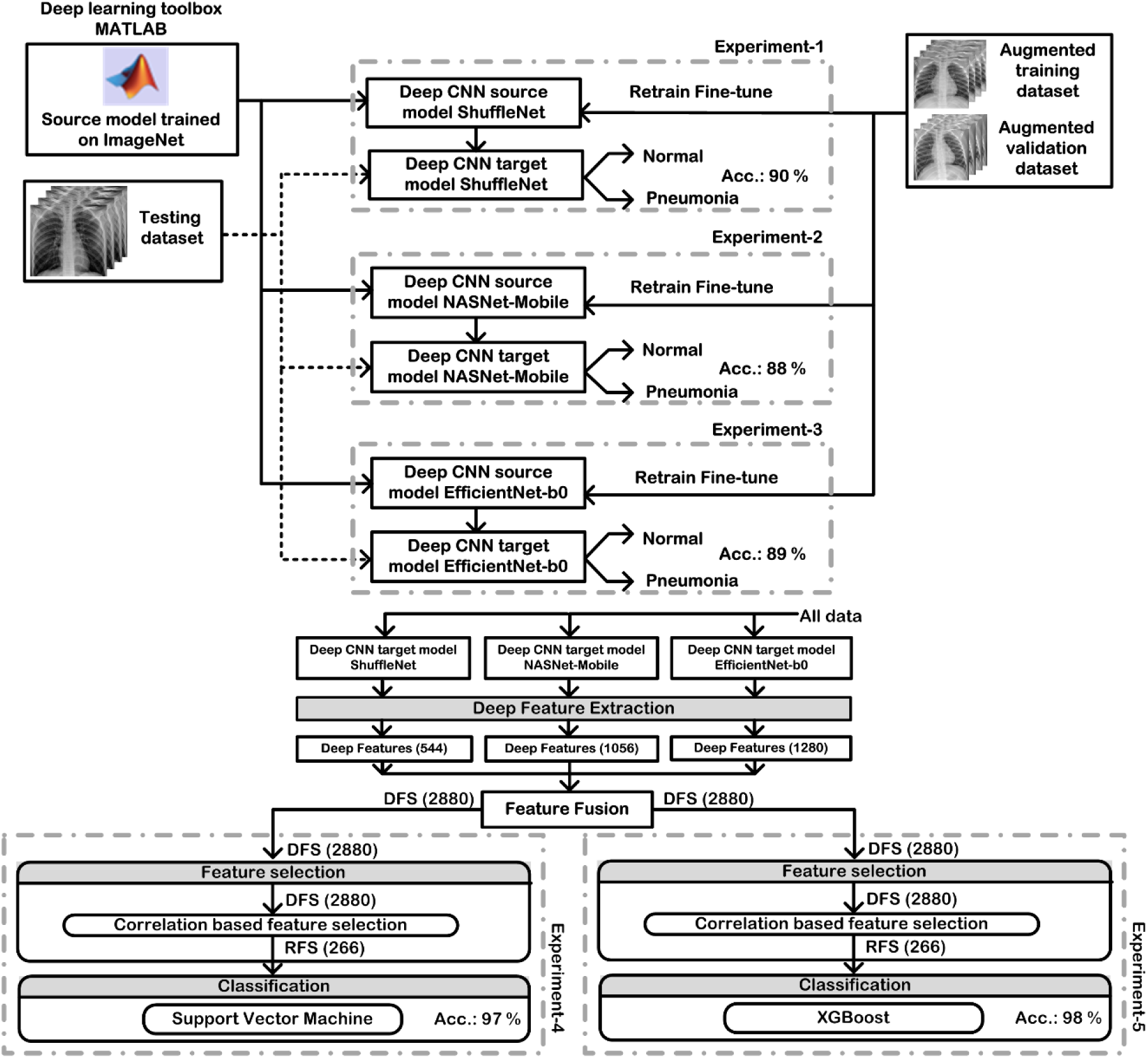
Experimental procedure used in proposed work.

The experiments conducted in the proposed work of lightweight CNN-based deep feature extraction, feature fusion CAD system design for chest radiographs are briefly described in Table 4.1.

**Table 4.1.**
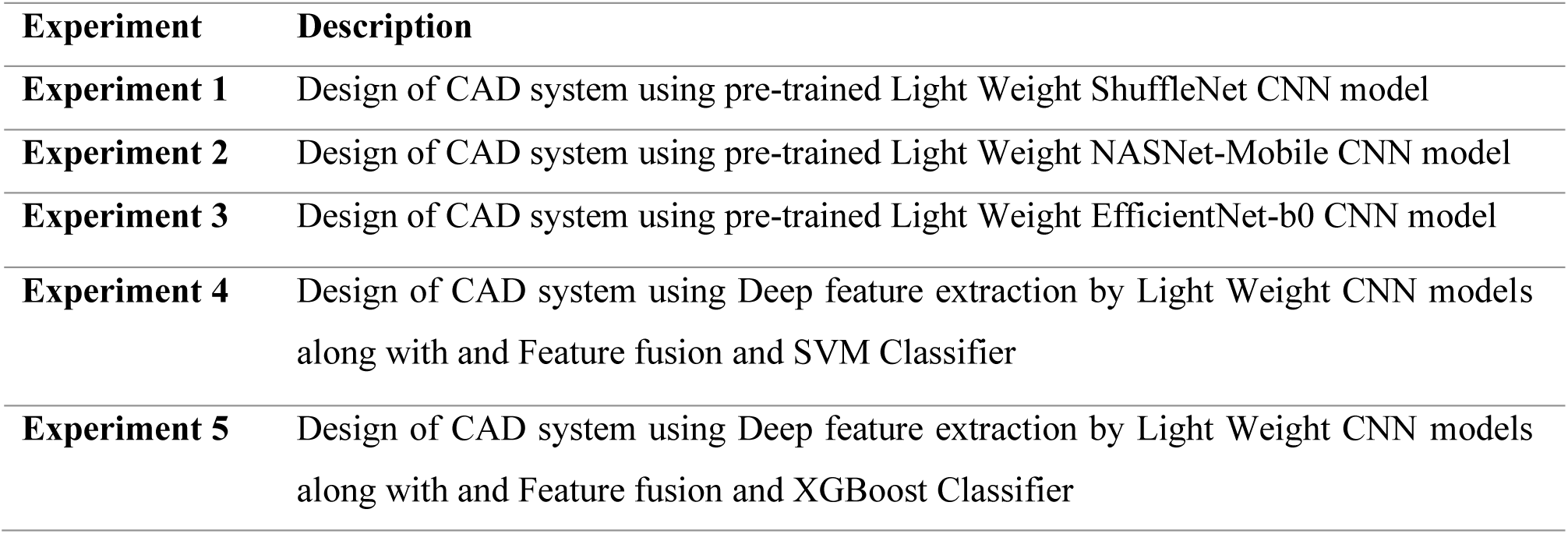
Overview of the Conducted Experiments in the Proposed Study.

### Experiment 1: Design of CAD system using pre-trained Light Weight ShuffleNet CNN model

This study focuses on developing a computer-aided diagnosis (CAD) system based on a pre-trained lightweight ShuffleNet CNN architecture. To improve classification accuracy, the model was trained on an augmented dataset of chest X-ray images. The performance of this CAD system, designed for binary classification of chest radiographs, is presented in Table 4.2.

**Table 4.2.**
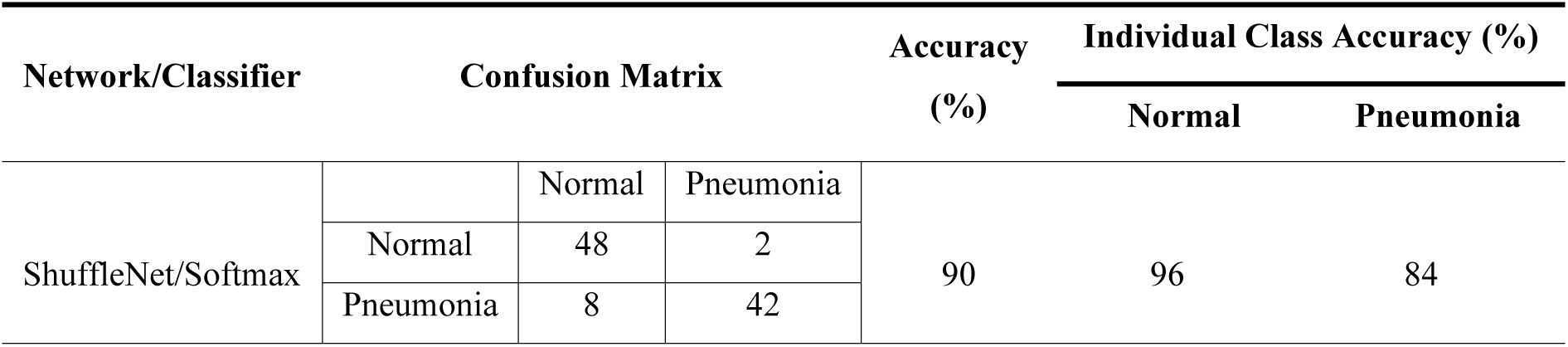
Performance evaluation of lightweight pre-trained ShuffleNet CNN model.

| Network/Classifier | Confusion Matrix |  |  | Accuracy (%) | Individual Class Accuracy (%) |  |
| --- | --- | --- | --- | --- | --- | --- |
|  |  |  |  |  | Normal | Pneumonia |
| ShuffleNet/Softmax |  | Normal | Pneumonia | 90 | 96 | 84 |
|  | Normal | 48 | 2 |  |  |  |
|  | Pneumonia | 8 | 42 |  |  |  |

### Experiment 2: Design of CAD system using pre-trained Light Weight NASNet-Mobile CNN model

This study focuses on developing a computer-aided diagnosis (CAD) system based on the pre-trained lightweight NASNet-Mobile convolutional neural network (CNN). To improve classification accuracy, the model was trained on an augmented dataset of chest X-ray images. The performance of this CAD system, designed for binary classification of chest radiographs, is presented in Table 4.3.

**Table 4.3.** Performance evaluation of lightweight pre-trained NASNet-Mobile CNN model.

| Network/Classifier | Confusion Matrix |  |  | Accuracy (%) | Individual Class Accuracy (%) |  |
| --- | --- | --- | --- | --- | --- | --- |
|  |  |  |  |  | Normal | Pneumonia |
| NASNet-Mobile /<br>Softmax |  | Normal | Pneumonia | 88 | 100 | 76 |
|  | Normal | 50 | 0 |  |  |  |
|  | Pneumonia | 12 | 38 |  |  |  |

### Experiment 3: Design of CAD system using pre-trained Light Weight EfficientNet-b0 CNN model

This study focuses on developing a computer-aided diagnosis (CAD) system based on the pre-trained lightweight EfficientNet-B0 convolutional neural network (CNN). The model was trained on an augmented dataset of chest X-ray images to improve classification accuracy. Table 4.4 presents the performance metrics of the CAD system in binary classification of chest radiographs using the EfficientNet-B0 architecture.

**Table 4.4.** Performance evaluation of lightweight pre-trained EfficientNet-b0 CNN model.

| Network/Classifier | Confusion Matrix |  |  | Accuracy (%) | Individual Class Accuracy (%) |  |
| --- | --- | --- | --- | --- | --- | --- |
|  |  |  |  |  | Normal | Pneumonia |
| EfficientNet-b0 /<br>Softmax |  | Normal | Pneumonia | 89 | 100 | 78 |
|  | Normal | 50 | 0 |  |  |  |
|  | Pneumonia | 11 | 39 |  |  |  |

### Experiment 4: Design of CAD system using Deep feature extraction by Light Weight CNN models along with Feature fusion and SVM Classifier.

This study focuses on developing a computer-aided diagnosis (CAD) system by applying deep feature extraction on the models trained from experiment 1 to 3. A total of 544 features from ShuffleNet, 1056 features from NASNet-Mobile and 1280 features from EfficientNet-b0 are extracted from the average pooling layer of each lighweight CNN model resulting in a deep feature set of a total of 2880 features i.e DFS = 2880, which further undergoes corelation based feature selection resulting in a reduced feature set of 266 features, i.e RFS = 266. Table 4.5 presents the performance evaluation results of the CAD system implemented using Deep feature extraction by Light Weight CNN models along with Feature fusion and SVM Classifier.

**Table 4.5.**
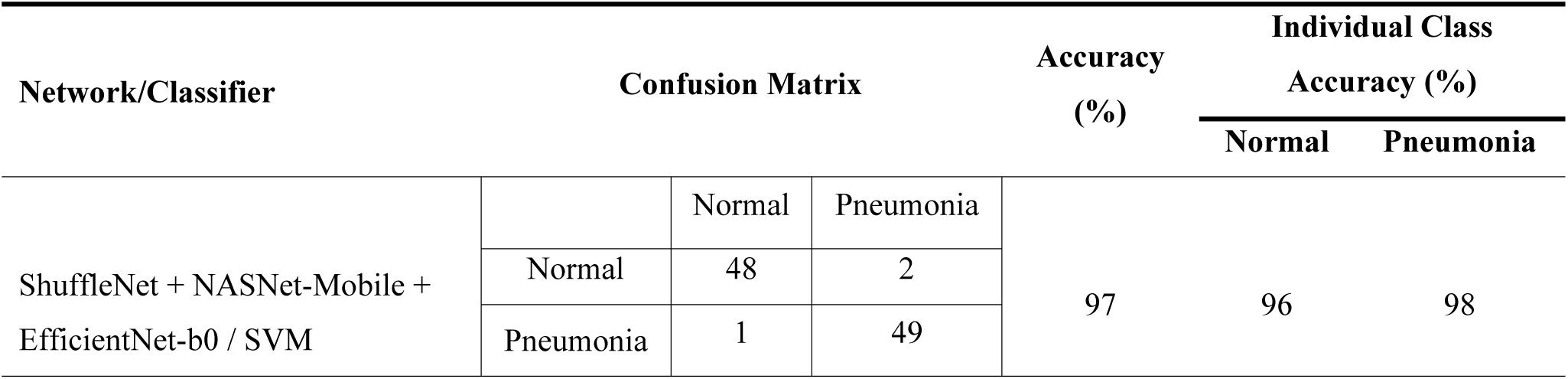
Performance Evaluation of CAD System Designed Using Deep feature extraction by Light Weight CNN models along with Feature fusion and SVM Classifier.

| Network/Classifier | Confusion Matrix |  |  | Accuracy (%) | Individual Class |  |
| --- | --- | --- | --- | --- | --- | --- |
|  |  |  |  |  | Accuracy (%) |  |
|  |  |  |  |  | Normal | Pneumonia |
| ShuffleNet + NASNet-Mobile + EfficientNet-b0 / SVM |  | Normal | Pneumonia | 97 | 96 | 98 |
|  | Normal | 48 | 2 |  |  |  |
|  | Pneumonia | 1 | 49 |  |  |  |

### Experiment 5: Design of CAD system using Deep feature extraction by Light Weight CNN models along with Feature fusion and XGBoost Classifier

This study focuses on developing a computer-aided diagnosis (CAD) system by applying deep feature extraction on the models trained from experiment 1 to 3. From each lightweight CNN models average pooling layer the features are extracted resulting in a DFS of 2880 features, which further undergoes corelation based feature selection resulting in a RFS of 266 features. The results of performance evaluation of the CAD system designed using Deep feature extraction by Light Weight CNN models along with Feature fusion and XGBoost Classifier is shown in Table 4.6.

**Table 4.6.**
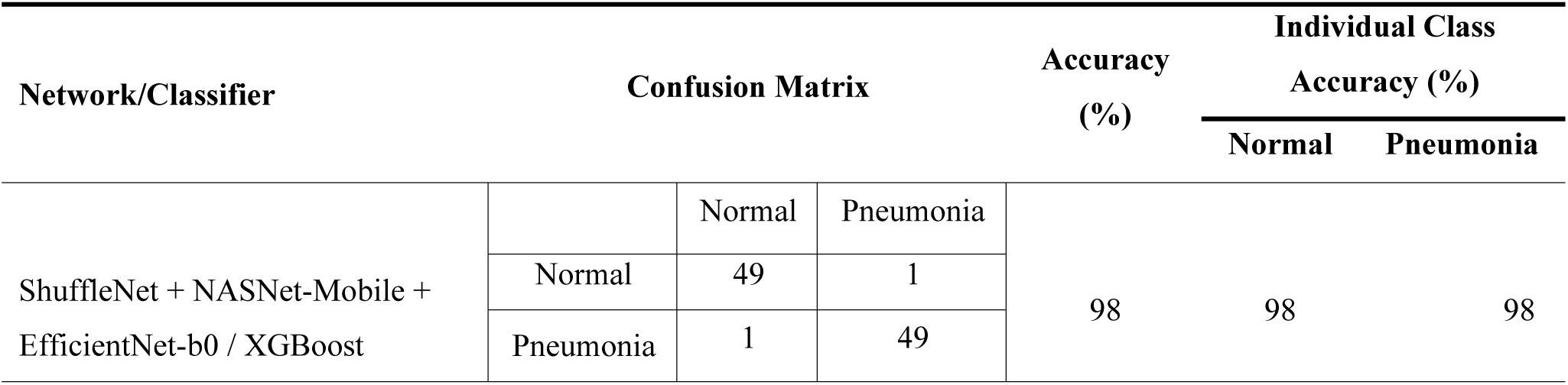
Performance Evaluation of CAD System Designed Using Deep feature extraction by Light Weight CNN models along with Feature fusion and XGBoost Classifier.

## 5. Result & Discussions

The comparative evaluation demonstrates that while individual lightweight CNN model ShuffleNet with 90% accuracy achieve respectable performance, the highest diagnostic efficacy is attained through feature fusion of multiple CNNs ShuffleNet + NASNet-Mobile + EfficientNet-b0 combined with XGBoost, achieving 98% balanced accuracy with 98% for both Normal and Pneumonia classes. This hybrid approach outperforms single-model CNNs and SVM-based fusion with 97% accuracy, highlighting that integrating complementary deep features with ensemble classifiers optimally balances sensitivity which deals with Pneumonia class detection and specificity which deals with Normal class identification, making it the most clinically reliable CAD system for chest X-ray classification.

The results emphasize the superiority of feature diversity and XGBoost’s discriminative power in medical image analysis. These results align with prior studies emphasizing the role of feature optimization in improving CAD system performance [101–115]. However, the current work advances the field by integrating lightweight CNNs with statistical feature selection and feature extraction, offering a balance between accuracy and efficiency as shown in Table 5.1.

**Table 5.1.** Comparative Evaluation of CAD System Implementations for Binary Classification of Chest X-ray Images.

| S. No. | Experiment | Network/Classifier | Accuracy (%) | ICA_Normal (%) | ICA_Pneumonia (%) |
| --- | --- | --- | --- | --- | --- |
| 1 | Design of CAD system using pre-trained Light Weight ShuffleNet CNN model | ShuffleNet/Softmax | 90.00 | 96.00 | 84.00 |
| 2 | Design of CAD system using pre-trained Light Weight NASNet-Mobile CNN model | NASNet-Mobile /Softmax | 88.00 | 100.00 | 76.00 |
| 3 | Design of CAD system using pre-trained Light Weight EfficientNet-b0 CNN model | EfficientNet-b0 /Softmax | 89.00 | 100.00 | 78.00 |
| 4 | Design of CAD system using Deep feature extraction by Light Weight CNN models along with Feature fusion and SVM Classifier | ShuffleNet + NASNet-Mobile + EfficientNet-b0/SVM | 97.00 | 96.00 | 98.00 |
| 5 | Design of CAD system using Deep feature extraction by Light Weight CNN models along with Feature fusion and XGBoost Classifier | ShuffleNet + NASNet-Mobile + EfficientNet-b0/XGBoost | 98.00 | 98.00 | 98.00 |

**Note:** ICA\_Normal: Individual Class Accuracy for Normal Class, ICA\_Pneumonia: Individual Class Accuracy for Pneumonia Class

The Fig. 5.1 illustrateds the ROC curve with its corresponding AUC values for the CAD system designed in the proposed work.

**Fig. 5.**
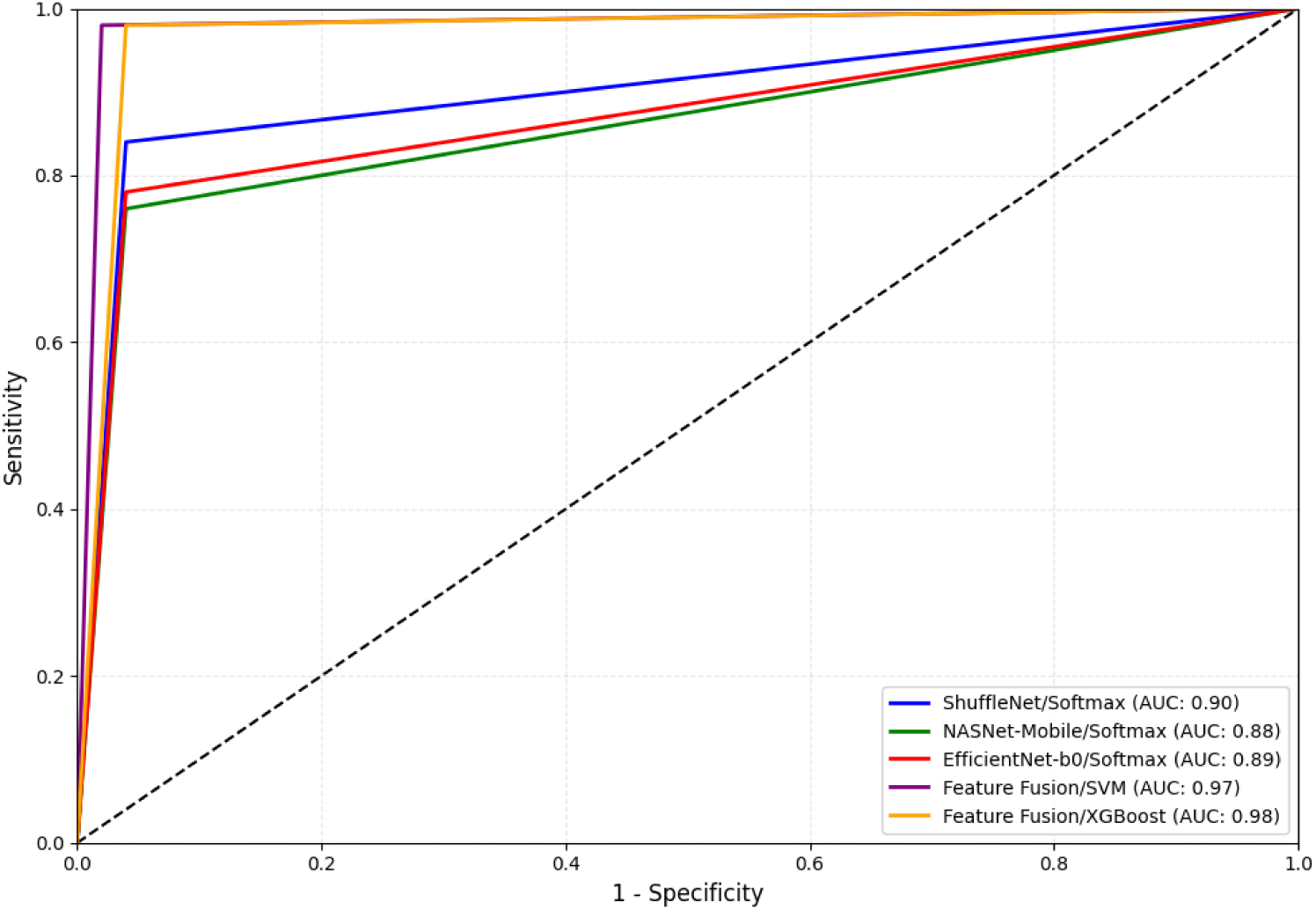
The ROC curve with its corresponding AUC values for the CAD system designed in the proposed work.

## 6. Conclusion

This study presented an efficient Computer-Aided Diagnosis system for pneumonia detection using chest X-ray images, leveraging the lightweight CNN models as deep feature extractors then further applying feature fusion and corelation based feature selection along with SVM classifier as well as XGBoost classifier. The experimental results demonstrated that the proposed model of CAD system using Deep feature extraction by Light Weight CNN models along with Feature fusion and XGBoost Classifier achieved 98% classification accuracy, highlighting its potential for accurate and rapid medical image analysis.

## Data Availability

All data produced are available online at https://www.kaggle.com/datasets/andrewmvd/pediatric-pneumonia-chest-xray

